# Characterizing the impact of the COVID-19 pandemic on HIV testing among Medicaid beneficiaries

**DOI:** 10.64898/2026.02.12.26346199

**Authors:** Maylin C. Palatino, Jacqueline E. Rudolph, Yiyi Zhou, Keri L. Calkins, Karine Yenokyan, Gregory M. Lucas, Xiaoqiang Xu, Eryka L. Wentz, Corinne E. Joshu, Bryan Lau

## Abstract

**Objectives:** Estimate the HIV testing, diagnoses, and test positivity rates among Medicaid beneficiaries in 2016-2021 and assess the impact of the COVID-19 pandemic on these outcomes.

**Design:** Prospective observational study of Medicaid enrollment, inpatient, and outpatient claims data from 27 states, 2016-2021.

**Methods:** We assessed Medicaid claims from adult beneficiaries with full benefits whose first continuous enrollment was ≥6 months without dual enrollment in other insurance, and without previous HIV diagnosis. We estimated the rates of annual testing, HIV diagnosis, and proportion of positive HIV tests among the tested using Poisson regression models. Bayesian structural time series modelling was performed to examine the pandemic’s impact on study outcomes with 3/16/2020-12/31/2021 as the pandemic period. We estimated rates overall and by age, sex, race/ethnicity, and states’ level of COVID-19-related restriction policies.

**Results:** We included 20,508,785 beneficiaries. Male beneficiaries, especially 18-34-year-olds, had lower annual testing uptake and higher test positivity rates than female beneficiaries. Black beneficiaries had higher annual testing rates than White and Hispanic beneficiaries. While the pandemic acutely disrupted the increasing pre-pandemic testing trend, the rates recovered to the expected level had the pandemic not happened, except among 18-34-year-old male beneficiaries, whose pandemic rates were, on average, 18.1% lower (95% confidence interval:-22.3,-13.8) than projected rates. HIV diagnosis and test positivity rates were not affected by the pandemic.

**Conclusion:** The pandemic significantly impacted the testing uptake among young male beneficiaries, highlighting the need for innovative strategies to improve HIV testing uptake in this demographic, restoring it to pre-pandemic levels or better.

## Introduction

The national emergency declaration in the United States (US) in March 2020 in response to the COVID-19 pandemic resulted in the disruption of routine, emergency, and non-emergency health care delivery^[1,2]^, including HIV testing^[3,4]^. The Center for Disease Prevention and Control (CDC) recommends that people ≥15 years old should be tested at least once in their lifetime and that those at high-risk for HIV acquisition should be tested annually^[5]^. Barriers to HIV testing access during the pandemic included HIV/sexually transmitted infection (STI) clinic closures, redeployment of HIV/STI staff to COVID-related activities, shelter-in-place and social distancing measures, interruption to public transportation, and delay or avoidance of medical care due to fear of COVID-19 exposure^[4,6–9]^. The interruption of HIV testing services during the pandemic could threaten the goal of reducing new HIV infections by ≥90% by 2030 as part of the Ending the HIV Epidemic in the US (EHE)^[10]^.

Several studies using clinic, outpatient, and/or inpatient data have shown a steep decline in HIV testing uptake in the US during the early pandemic^[11–17]^. This decline was followed by an increasing trend in the second half of 2020; however, results have varied on whether testing uptake returned to the 2019 level^[11–18]^. Moreover, the impact of the pandemic on HIV testing across various demographic groups remains unclear, underscoring the importance of identifying these effects to formulate targeted HIV prevention policies that protect those most vulnerable to disruptions in health services. Furthermore, because most prior studies focused on data through 2020, estimating HIV testing and test positivity rates beyond 2020 would clarify whether these rates returned to pre-pandemic levels as the health system recovers.

This paper aims to estimate the rates of HIV testing, HIV incidence, and test positivity among Medicaid beneficiaries from 2016 to 2021 and to quantify the impact of the COVID-19 pandemic on these outcomes overall and by demographic characteristics. Medicaid is the primary source of health coverage for individuals who are low income or have disabilities, insuring 15% of the US adult population between 2016 and 2021^[19]^. In addition to filling in the gap on HIV testing uptake overall during the pre-pandemic and pandemic periods, the diversity and large sample size of Medicaid beneficiaries allow for stratified estimation of all three HIV outcomes (testing, incidence, test positivity).

## Methods

### Study Sample

We utilized enrollment, inpatient, outpatient, and long-term care claims data from the Centers for Medicare and Medicaid Services of Medicaid beneficiaries from the District of Columbia (DC) and 26 US states (see Supplementary Fig. 1 for the map of included states). We included beneficiaries aged 18-64 years whose first continuous enrollment in Medicaid between 2016 and 2021 was >6 months, had full benefits, without dual enrollment in Medicare or private insurance, and without previous HIV diagnosis. HIV diagnoses were based on one inpatient or two outpatient claims with HIV-related ICD-9/ICD-10 code within two years of each other^[20,21]^ (see Supplementary Table 1 for codes). Analytic baseline was defined as six months after the start of the first eligible Medicaid enrollment period. Eligible beneficiaries were followed until HIV diagnosis, Medicaid disenrollment, 65^th^ birthday, death, or 12/31/2021, whichever was earliest. This secondary analysis of existing Medicaid claims data was determined to meet the criteria for exemption by the Johns Hopkins Bloomberg School of Public Health Institutional Review Board.

### Measures

The study outcomes were HIV testing, diagnosis (defined above), and test positivity. HIV testing was based on one outpatient or inpatient claim using related ICD-10 and CPT/HCPCS codes (Supplementary Table 1). To address possible repeat testing from inconclusive results, only the last test within any 30-day period was counted per beneficiary. For test positivity, we considered an HIV test to be positive if it was the latest HIV test claim for a beneficiary and was followed by an HIV diagnosis within 30 days. In a sensitivity analysis, we considered diagnoses occurring within 60 days of a positive test. We obtained demographic and enrollment information from personal summary file, including sex, race/ethnicity (non-Hispanic White [White hereafter], non-Hispanic Black [Black hereafter], Hispanic, other), US state, birth date, death date, Medicaid enrollment date, and disenrollment date. In accordance with a previously published definition from the Pew Research Center^[22]^, states in our analysis were grouped and classified as most, less, or least restrictive (Supplementary Fig. 1) based on the number of preventive measures implemented per state around the first week of November 2020 (e.g., stay-at-home orders, mandatory face coverings, closures of non-essential businesses). Although the Pew Research Center utilized restriction policies from early-November 2020, we believe these groupings accurately represent the variations in states’ COVID-19 prevention policies throughout the pandemic. States in our analysis were also classified by US Census region (Supplementary Fig. 1).

### Statistical analysis

To determine the estimated annual rate of HIV testing, diagnosis, and test positivity, the data were divided into yearly periods. In each year a beneficiary was enrolled, they contributed person-time from the start of the year or their baseline (if enrolling mid-year), whichever was earlier, to the earlier of the end of the year or end of follow-up (if it ended mid-year). An indicator variable for HIV testing was created and coded as 1 if a beneficiary had ≥1 test dates in a specific year or 0 otherwise. The crude incidence rates of HIV testing and HIV diagnosis per year were computed as the number of tests and the number of HIV diagnoses within that year, respectively, divided by the sum of the person-year contribution of each beneficiary within the year multiplied by 10,000 for diagnosis or 100 for testing. The annual crude test positivity rate was estimated as the proportion of the number of beneficiaries with positive tests out of the number that were tested in the year. To estimate adjusted rates, Poisson regression modelling was employed including age at baseline (specified as natural cubic splines), sex, race/ethnicity, and US state as covariates. For each outcome, the annual adjusted rate was the mean of the predicted probabilities of having the outcome. The 95% confidence intervals (CI) of the estimates were the 2.5^th^ and 97.5^th^ percentiles of the point estimates from 1000 bootstrap resamples.

To quantify the impact of the COVID-19 pandemic on HIV testing, diagnosis, and test positivity rates, we included data from January 1, 2017, to December 31, 2021, where the pre-pandemic and pandemic periods were before and on or after March 16, 2020, respectively. We removed 2016 in this analysis as data from 2017 to 2021 provided more stable prediction estimates for the Bayesian structural time series (BSTS). The data were divided into bimonthly periods where the 1^st^ to the 15^th^ day and the 16^th^ to the last day of the month were the first and second period per month, respectively. The person-time contribution of a beneficiary and the crude rates were defined as described above, just on the bimonthly scale instead of annual. We used direct standardization, following the method described by Naing,^[23]^ to standardize the bimonthly rates of each outcome to the distribution of age, sex, race/ethnicity, and region observed at the intervention time (mid-March 2020). We then performed BSTS analysis using the standardized rates^[24–26]^.

Briefly, a BSTS model is a random state-space model that presents the trends, seasonality, and regression components separately. It combines prior information with the likelihood function to produce the posterior distribution. The forecasts using the BSTS model are obtained from the sample results of the posterior distribution using the Markov Chain Monte Carlo (MCMC) algorithm. We used the *bsts* R package^[27]^ to build the BSTS models with the MCMC algorithm with 10,000 iterations and burn-in of 2,000 to ensure sufficient samples and convergence. We then used the *CausalImpact* package^[24]^ to estimate the impact of the COVID-19 pandemic on the outcome variables using the built BSTS model in a manner analogous to an interrupted time series model and reported the relative effects (RE) and 95% credible intervals (CrI). The RE was the difference between the average predicted value and the average observed value during the pandemic period divided by the average predicted value in percent. We employed the *ggplot2* R package^[28]^ to create the figures in the paper. We carried out analyses overall and stratified by age, sex, race/ethnicity, and states’ level of COVID-19-related restriction policies.

## Results

We included 20,508,785 beneficiaries with a median follow-up time of 1.5 years [interquartile range (IQR):0.6,3.2] (Supplementary Table 2). The majority were female (54.8%), and the median baseline age was 26.7 years (IQR:18.5,40.7). Beneficiaries were predominantly White (44.0%), followed by other races/ethnicities (32.9%) and Black (19.9%). The three largest contributing states were California (15.9%), Michigan (9.0%), and Kentucky (6.4%) (Table 1).

**Table 1.**
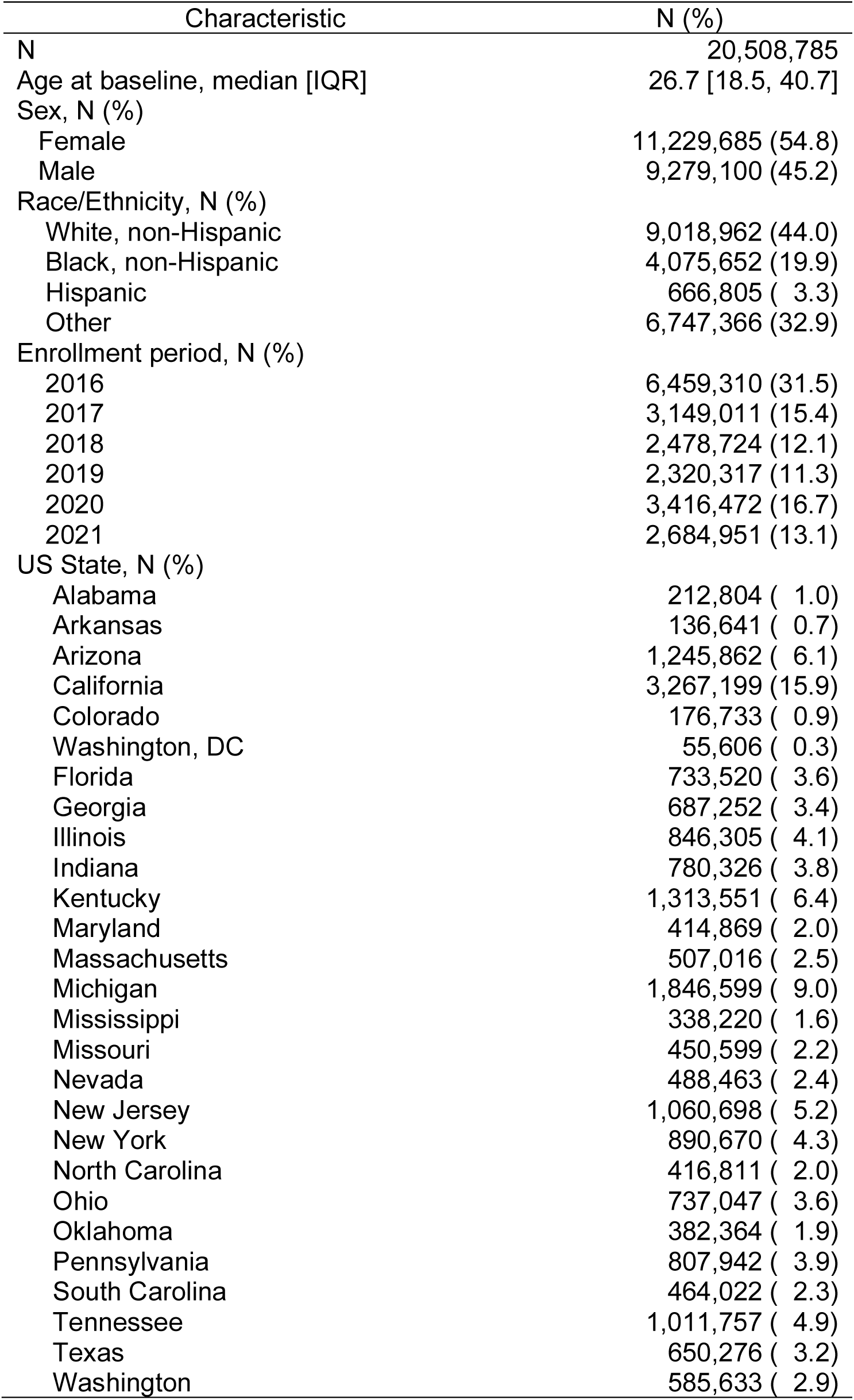
Baseline characteristics of the included Medicaid beneficiaries.

### HIV testing

#### Annual incidence rate of HIV testing

The overall annual adjusted testing rates (per 100 person-years) increased from 2016 to 2019 [2016: 3.9 (95%CI:3.9,3.9), 2017: 6.7 (95%CI:6.7,6.7), 2018: 7.1 (95%CI:7.0,7.1), 2019: 7.6 (95%CI:7.5,7.6)], dropped to 6.8 (95%CI:6.7,6.8) in 2020, and increased to 8.4 (95%CI:8.4,8.4) in 2021 (Table 2).

**Table 2.**
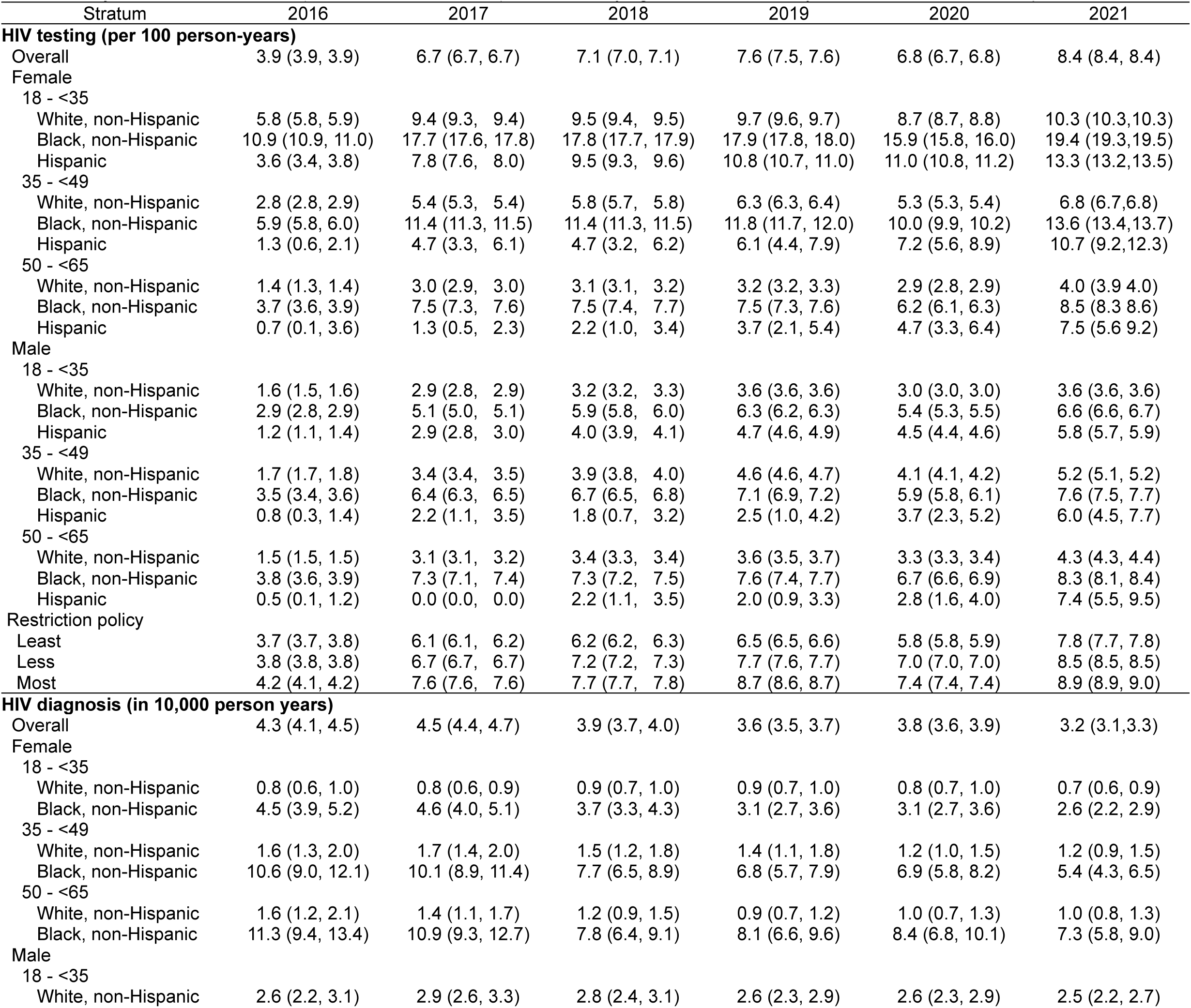
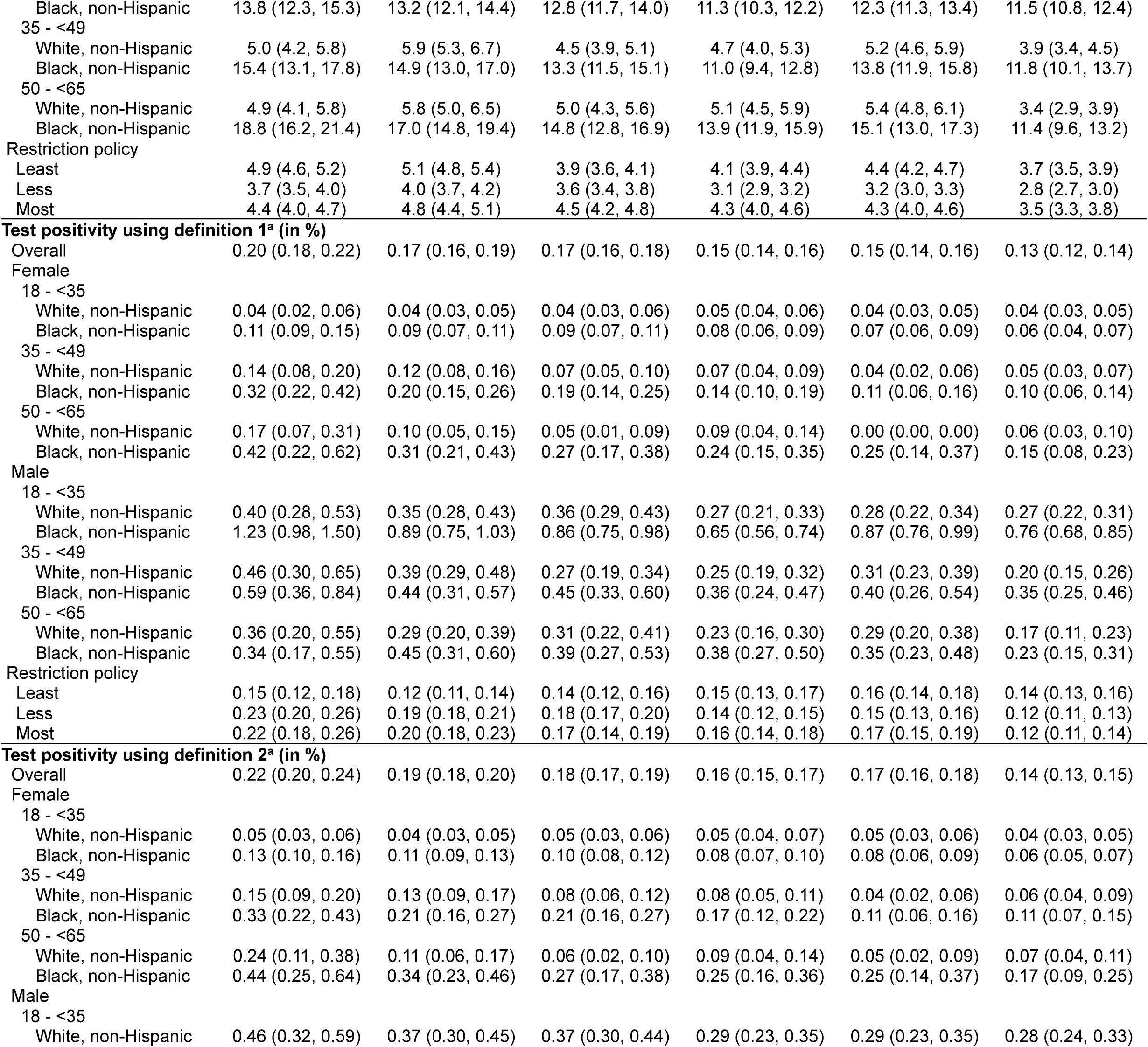
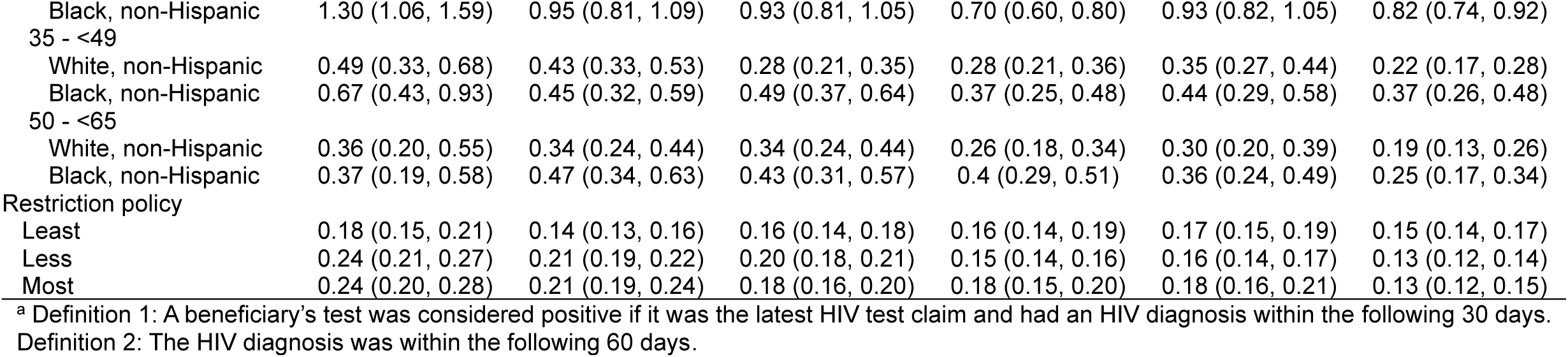
Adjusted annual estimates (95% confidence interval) overall and by age, sex, race/ethnicity, and COVID-19 restriction policies.

Male beneficiaries aged 18-34 had lower race/ethnicity-specific annual HIV testing rates compared to 18-34-year-old female beneficiaries (Fig. 1, Table 2). The same was observed among Black and White 35-49-year-old male beneficiaries compared to female beneficiaries in the same age group. Black beneficiaries had higher annual rates compared to White and Hispanic beneficiaries, for most age groups and sex. Overall, the age-specific testing rates of male and female Hispanic beneficiaries had an increasing trend across years.

**Figure 1.**
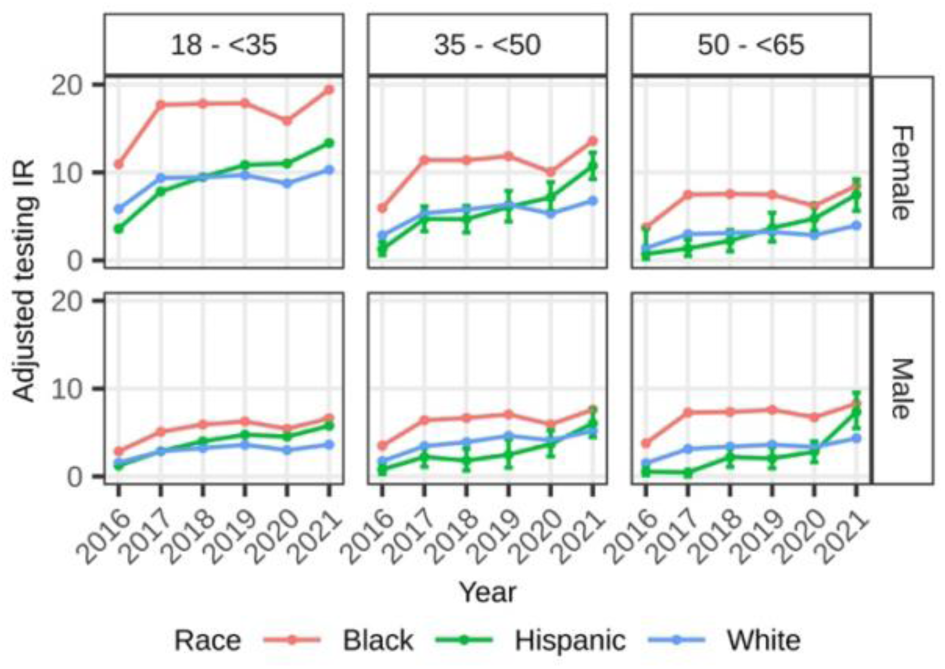
Adjusted annual HIV testing incidence rate among Medicaid beneficiaries without HIV by age, sex and race/ethnicity, 2016-2021

Stratifying by states’ level of COVID-19 restriction policies, states with the strictest policies had the highest annual testing rates from 2016-2021 compared to states with the least and less restrictive policies. The annual testing rates were lowest among states with the least restrictive policies from 2017-2021 (Table 2). The testing rates for all categories dropped in 2020, with states with the strictest policies having the steepest drop.

#### Effect of COVID-19 pandemic on HIV testing

We saw a sharp decline in the standardized testing rate in the first three months of the pandemic, followed by recovery starting July 2020 (Fig. 2A). The difference in the predicted and observed rates was largest in the early pandemic period. For most of the pandemic period, the observed values were lower than the predicted values but within the 95% CrI. On average, the observed standardized HIV incidence rates in logarithmic scale were lower by 8.1% (95%CrI:-12.5,-3.4) than can be expected had the pandemic not happened (see Supplementary Table 3 for the RE overall and by strata).

**Figure 2.**
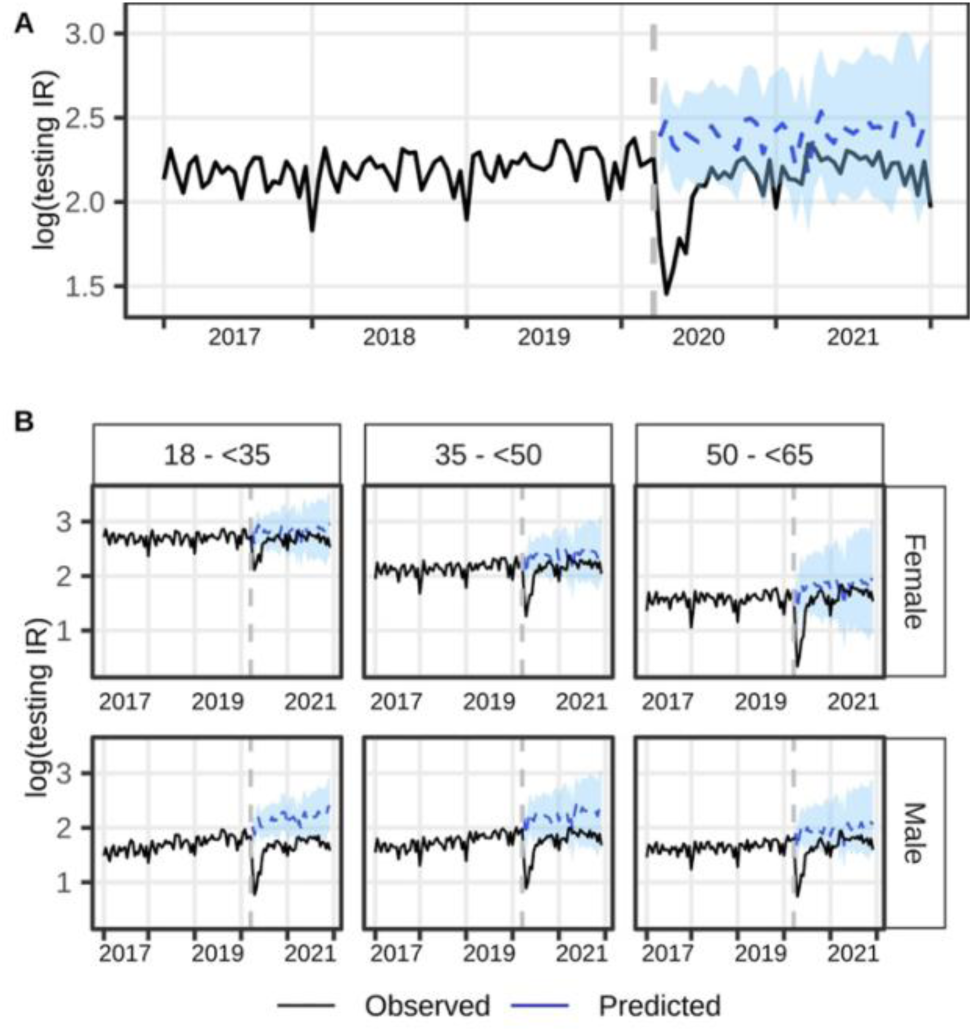
Trend of log(standardized HIV testing incidence rate) from January 2017 to December 2021 A). overall, and B) stratified by age and sex. The vertical gray dashed line separated the pre-pandemic and pandemic periods. The solid lines represent the observed values while the blue dashed lines with blue 95% credible intervals during the pandemic period represent the predicted values had the pandemic not happened.

All age-sex-specific standardized testing rates during the pandemic period were lower as compared to what would have been expected had the pandemic not happened (Fig. 2B, Supplementary Table 3). Male beneficiaries aged 18-34 (RE=-18.1; 95%CrI:-22.3,-13.8) and 35-49 (RE=-14.6; 95%CrI:-20.6,-7.3) had the largest observed-expected rate difference, while 18-34-year-old female beneficiaries had the smallest observed-expected rate difference (RE=-5.1; 95%CrI:-9.2,-0.3). Although we observed a steep decline in the testing incidence rates during the initial months of the pandemic followed by a subsequent increase in all age-sex strata, the observed rates were within the 95% CrI of the predicted values by July 2020 among female beneficiaries across all age strata, and by September 2020 among 35-49- and 50-64-year-old male beneficiaries. The observed testing rates of 18-34-year-old male beneficiaries were significantly lower than the predicted rates for most of the pandemic period (Fig. 2B).

When examining age-race/ethnicity-specific rates, Black beneficiaries ≥50 years old (RE=-12.2; 95%CrI:-23.8, −0.5) and 35-49-year-old White beneficiaries (RE=-12.2; 95%CrI:-18.3, −3.6) had the highest observed-expected rate difference, while 18-34-year-old Black beneficiaries (RE=-5.3; 95%CrI:-9.3, −0.3) had the lowest difference (Supplementary Table 3). Despite the dip in the early pandemic period in all age-race/ethnicity and restriction strata (Supplementary Fig. 2), the rates were not statistically different from the predicted rates starting June 2020 until the end of 2021.

### HIV incidence

#### Annual incidence rate of HIV diagnoses

The adjusted HIV incidence rates (per 10,000 person-years) decreased from 2017 (4.5, 95%CI:4.4,4.7) to 2018 (3.9, 95%CI:3.7,4.0) and remained stable from 2019 to 2021 [2019: 3.6 (95%CI:3.5,3.7), 2020: 3.8 (95%CI:3.6,3.9), 2021: 3.2 (95%CI:3.1,3.3)] (Table 2). The age-sex-race/ethnicity-specific HIV incidence rates among Black beneficiaries across years were higher compared to White beneficiaries (Fig. 3A). For each age category, the incidence rates were higher among male than female beneficiaries, especially among Black beneficiaries. Overall, there was a decreasing trend in the yearly estimates among Black female beneficiaries in all age strata and among Black males in the oldest age group, such that the 2021 estimates were significantly lower than the 2017 estimates. This was not observed among 18-49-year-old Black male beneficiaries. Rates for Hispanic beneficiaries were not estimated due to sparse events.

**Figure 3.**
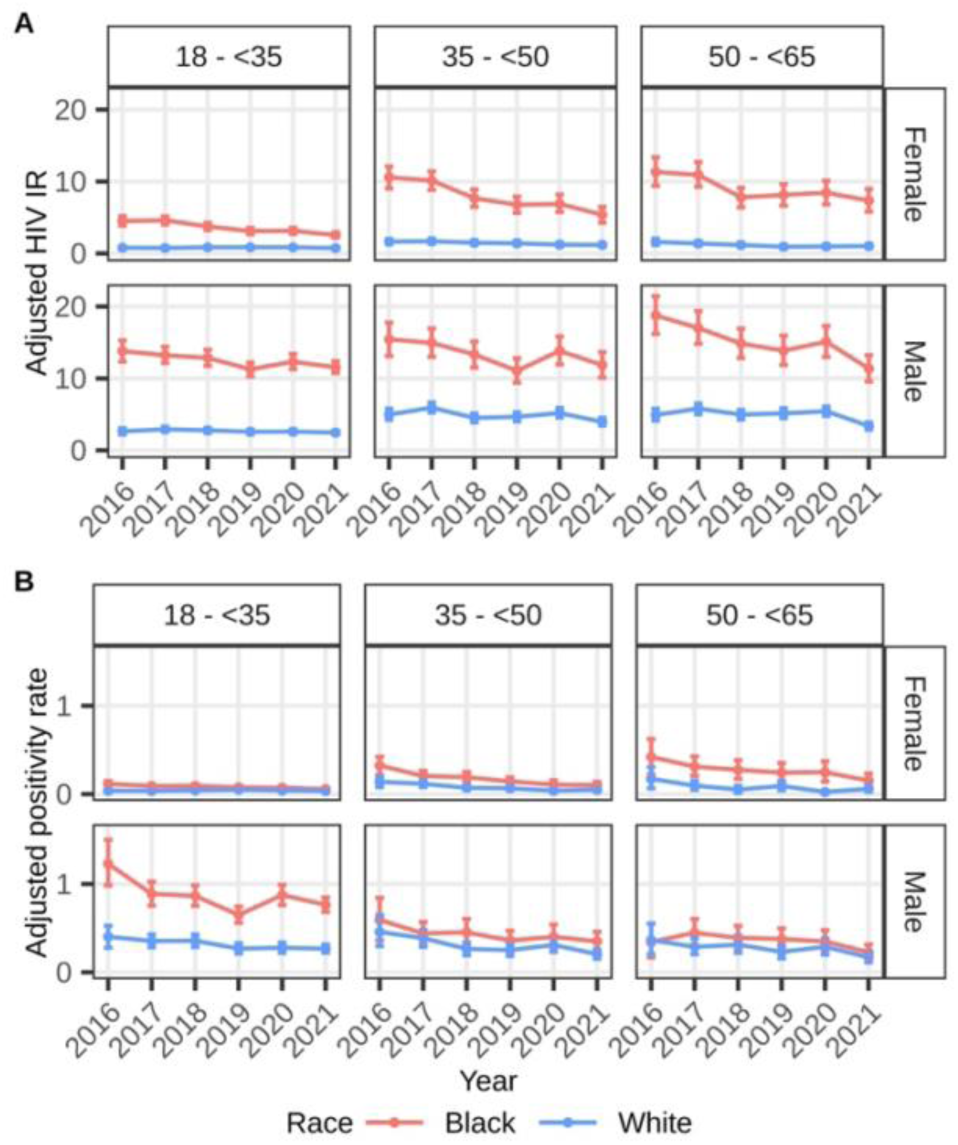
A) Adjusted HIV incidence rates (per 10,000 person-years) and B) adjusted annual test positivity rates (in %) among Medicaid beneficiaries without HIV by age, sex, and race/ethnicity, 2016 – 2021

The annual rates for states with the less restrictive policies were lower than those with the most and least restrictive policies for all years except in 2018. States with the most and least restrictive policies had comparable rates per year except in 2018 (Table 2).

#### Effect of COVID-19 pandemic on HIV incidence

The observed incidence rates of HIV diagnoses during the pandemic were not statistically different from the expected values had the pandemic not happened, both overall and in all strata (Supplementary Fig. 3).

### Test positivity

#### Annual test positivity rate (in %)

The annual adjusted test positivity rates slightly declined across years [2016: 0.20 (95%CI:0.18,0.22), 2017: 0.17 (95%CI:0.16,0.19), 2018: 0.17 (95%CI:0.16,0.18), 2019: 0.15 (95%CI:0.14,0.16), 2020: 0.15 (95%CI:0.14,0.16), 2021: 0.13 (95%CI:0.12,0.14)]. A similar trend was observed when a positive HIV test for a beneficiary was defined as the latest HIV test claim followed by an HIV diagnosis within 60 days, rather than within 30 days that was set for the primary analyses (Table 2).

Black male beneficiaries aged 18-34 had the highest test positivity rates across years compared to other age-race/ethnicity-sex strata. This subgroup had a sharp increase in test positivity in 2020. White female beneficiaries had the lowest rates per age group of any race/ethnicity-sex groups (Fig. 3B, Table 2).

The adjusted test positivity rates in states with the most and less restrictive COVID-19 policies had a decreasing trend from 2016 to 2021, such that the 2021 estimates were significantly lower than the 2016 estimates. The states with the least restrictive policies had stable test positivity rates across years (Table 2).

#### Effect of COVID-19 pandemic on test positivity rate

The observed test positivity rates during the pandemic were not statistically different from the expected rates had the pandemic not happened, overall or stratified by sex, race, and COVID restriction policies (Supplementary Fig. 4).

## Discussion

We found that although the increasing trend in the HIV testing rates among Medicaid beneficiaries was disrupted during the first months of the pandemic, the overall HIV testing rates recovered and were not statistically different from the expected level had the pandemic not happened. This observation was seen for all age-sex and age-race/ethnicity strata except for 18-34-year-old male beneficiaries, whose HIV testing uptake remained lower than what was expected. Moreover, male beneficiaries consistently had lower annual testing uptake than female beneficiaries, and 18-34-year-old Black male beneficiaries had the highest annual test positivity rates. In contrast, 18-34-year-old Black female beneficiaries had the highest annual testing uptake, and an increasing testing uptake trend was observed among male and female Hispanic beneficiaries.

Our finding that the HIV testing rates were increasing overall from 2016-2019 aligns with the results of previous studies among Medicaid and privately insured individuals. A 2021 study investigating HIV testing uptake in 2014-2019 among Medicaid beneficiaries found similar, albeit slightly lower, HIV testing rates in 2016-2019^[29]^. This study included beneficiaries aged ≥13 years from all states, while we only included 18-64-year-old beneficiaries from 27 states, which could explain the differences in estimated rates.

Another study from Kaiser Permanente Southern California (KPSC) found a similar increasing trend in 2017-2019^[13]^. The said study had slightly higher testing rates than ours, which may be because the former included a more socioeconomically diverse KPSC members who were ≥12 years old.

During the early pandemic period, our study found a sharp decrease in testing uptake, while HIV diagnoses remained stable and the observed test positivity rates, albeit not statistically significant, were higher than the point estimates of the predicted rates. This suggests that, although overall testing declined, high-risk beneficiaries continued to be tested, leading to stable diagnoses rates and a slight rise in positivity rates.

The observed decline in HIV testing rates in the early pandemic periods, which may be attributed to stay-at-home orders and changes in service delivery^[12,13]^, aligns with findings of previous studies in various settings^[11,12,14–17]^. Our results further showed that the testing rates recovered after the initial decline and were not statistically different from the expected rates had the pandemic not happened in most groups. Previous studies using laboratory and electronic health records had varying results as regards testing rate recovery. A study done in four large, geographically diverse, urban-based healthcare systems^[12]^, one utilizing data from Oregon State Public Health Laboratory^[17]^, a CDC report covering four national data collection systems ^14^, and a study on CDC-funded HIV tests^15^ saw after the initial dip that the number of tests increased for the rest of 2020, although remained lower than 2019 level. Meanwhile, a study conducted in a large gender-minority-focused federally qualified health center in Chicago, Illinois^[16]^ and another using data from large commercial laboratories^[18]^ reported that the number of HIV tests rebounded to the 2019 level. Our study is the first to use claims data in assessing the impact of the pandemic on testing uptake.

Male beneficiaries had lower testing uptake from 2016-2021 as compared to female beneficiaries. We expected this result since we included pregnant women, who were recommended to have HIV screening^[30]^. Nonetheless, a previous study on Medicaid beneficiaries showed similar results in 2019^[29]^. This is alarming given that young men, especially those who report male-to-male sexual contact, comprise a large proportion of new HIV infections^[31]^. Low testing uptake coupled with high positivity rate signify potential for undiagnosed HIV in this demographic. Moreover, the HIV testing uptake of 18-34-year-old male beneficiaries did not recover even after two years into the pandemic, presenting possible onward transmission of undiagnosed infection and dire consequences for EHE efforts.

We likewise examined the outcome trends by states’ level of COVID-19 restriction policies. The states with the least strict restriction policies had the lowest HIV testing uptake in 2016-2021, while states with the strictest restrictions had the highest testing uptake for most years. These findings may be linked to Medicaid expansion implementation, which was found to be associated with an increase in HIV testing uptake^[32,33]^. States with the strictest restriction policies were among the first states to implement Medicaid expansion. Most states with the least restrictive policies did not adopt Medicaid expansion or only adopted in Mid-2021^[34]^. The drop in the 2020 testing rates, especially in the early pandemic period, was steepest in states with the strictest policies, which align with prior findings. World regions with more stringent lockdowns experienced a larger decrease in HIV testing during the pandemic than regions with less^[35]^. Despite this, our analyses showed recovery to the expected level had the pandemic not happened right after the early pandemic period for all levels of restriction policies.

This study had several limitations. First, claims data do not capture testing that was not financed through Medicaid, such as those done through HIV outreach events or STI clinics, which may result in underestimation of testing rates. Second, we only included beneficiaries who had ≥6 months of continuous enrollment, thus, possibly excluding those who were experiencing ‘churn’ (i.e., temporary loss of Medicaid coverage due to various reasons), particularly for pre-pandemic period coverage. During the pandemic period, the Family First Coronavirus Response Act prohibited states to reevaluate the Medicaid eligibility of beneficiaries until March 2023, thus, giving beneficiaries continuous coverage^[36]^. Experiences of ‘churn’ vary by demographic characteristics^[37]^, which could result in under- or overestimation of stratified estimates. Third, while we have procedure codes for HIV testing, test results are not available in claims data. We defined HIV diagnosis using diagnosis codes. HIV diagnosis codes entered within 30 or 60 days of the HIV testing date may not correspond to the claimed test, thus, overestimating test positivity estimates.

Our study is the first to examine the impact of the COVID-19 pandemic on the HIV testing uptake of Medicaid beneficiaries, a vulnerable population making up 15% of adult Americans^[19]^. The large sample size and varied demographic characteristics of beneficiaries allowed us to estimate stratified HIV testing incidence and other outcomes and to illustrate how the pandemic affected these outcomes. We found that, after the drop at the early pandemic period, the testing uptake recovered to the level comparable to the rate trajectory without the disruption of the pandemic. This was seen for all demographics except 18-34-year-old male beneficiaries, who had the lowest rates of testing across the study period and whose pre-pandemic increasing trend was disrupted and did not recover even until the end of 2021. This finding suggests the vulnerability of young male beneficiaries’ healthcare engagement to disruption in services and highlights the need for innovative strategies and interventions to increase HIV testing uptake in this demographic. Future studies may examine the annual incidence of multiple testing and the testing uptake of beneficiaries with high-risk sexual behavior diagnoses.

## Author Contributions

Study concept and design: Bryan Lau, Corinne Joshu

Data processing and analysis: Maylin Palatino, Xiaoqiang Xu

Interpretation of data: Maylin Palatino, Jacqueline Rudolph, Yiyi Zhou, Karine Yenokyan, Eryka Saylor, Gregory Lucas, Keri Calkins, Corrine Joshu, Bryan Lau.

Drafting of the manuscript: Maylin Palatino

Critical revision for important intellectual content: Maylin Palatino, Jacqueline Rudolph, Yiyi Zhou, Karine Yenokyan, Eryka Saylor, Gregory Lucas, Keri Calkins, Corrine Joshu, Bryan Lau.

All authors approved the final version of the manuscript.

## Data Availability

The data that support the findings of this study are under the authority of the Centers for Medicare & Medicaid Services (CMS) and administered by ResDAC. Investigators may reuse these data if they independently meet CMS requirements and obtain both CMS reuse approval and permission from the studys NIH program officer.

## References

1. Hartnett KP, Kite-Powell A, DeVies J, Coletta MA, Boehmer TK, Adjemian J, et al. Impact of the COVID-19 Pandemic on Emergency Department Visits — United States, January 1, 2019–May 30, 2020. MMWR Morb Mortal Wkly Rep 2020; 69:699–704.

2. Lange SJ, Ritchey MD, Goodman AB, Dias T, Twentyman E, Fuld J, et al. Potential Indirect Effects of the COVID-19 Pandemic on Use of Emergency Departments for Acute Life-Threatening Conditions-United States, January-May 2020. MMWR Morb Mortal Wkly Rep 2020; 69:795–800.

3. Simoncini GM, Armon C, Buchacz K, Mahnken J, Qingjiang H, Chagaris K, et al. STI Testing and Rates of STI Diagnoses Before and During the COVID-19 Pandemic in a US HIV Cohort. Sex Transm Dis. 2025; 52:304–309.

4. Santos GM, Ackerman B, Rao A, Wallach S, Ayala G, Lamontage E, et al. Economic, Mental Health, HIV Prevention and HIV Treatment Impacts of COVID-19 and the COVID-19 Response on a Global Sample of Cisgender Gay Men and Other Men Who Have Sex with Men. AIDS Behav. 2021; 25:311–321.

5. Dinenno EA, Prejean J, Irwin K, Delanet KP, Bowles K, Martin T, et al. Recommendations for HIV Screening of Gay, Bisexual, and Other Men Who Have Sex with Men — United States, 2017. MMWR Morb Mortal Wkly Rep 2017; 66:830–832.

6. Czeisler MÉ, Marynak K, Clarke KEN, Salah Z, Shakya I, Thierry JM, et al. Delay or Avoidance of Medical Care Because of COVID-19–Related Concerns — United States, June 2020. MMWR Morb Mortal Wkly Rep 2020; 69:1250–1257.

7. Darcis G, Vaira D. Moutschen M. Impact of coronavirus pandemic and containment measures on HIV diagnosis. Epidemiol Infect. 2020; 148:e185.

8. Ogunbodede OT, Zablotska-Manos I, Lewis DA. Potential and demonstrated impacts of the COVID-19 pandemic on sexually transmissible infections. Curr Opin Infect Dis 2021; 34:56–61. doi: 10.1097/QCO.0000000000000699.

9. Zapata JP, Dang M, Quinn KG, Horvath KJ, Stephenson R, Dickson-Gomez J, et al. COVID-19-Related Disruptions to HIV Testing and Prevention Among Young Sexual Minority Men 17–24 Years Old: A Qualitative Study Using Synchronous Online Focus Groups, April–September 2020. Arch Sex Behav 2022; 51:303–314. doi: 10.1007/s10508-021-02166-7

10. Center for Disease Control and Prevention. Ending the HIV Epidemic in the US Goals. https://www.cdc.gov/ehe/php/about/goals.html. [Accessed 10 October 2025]

11. Hill BJ, Anderson B, Lock L. COVID-19 Pandemic, Pre-exposure Prophylaxis (PrEP) Care, and HIV/STI Testing Among Patients Receiving Care in Three HIV Epidemic Priority States. AIDS Behav. 2021; 25:1361–1365.

12. Moitra E, Tao J, Olsen J, Shearer RD, Wood BR, Busch AM, et al. Impact of the COVID-19 pandemic on HIV testing rates across four geographically diverse urban centres in the United States: An observational study. Lancet Reg Health Am 2022;7: 100159 (2022).

13. Chang JJ, Chen Q, Dionne-Odom J, Hechter RC, Bruxvoort, KJ. Changes in Testing and Diagnoses of Sexually Transmitted Infections and HIV During the COVID-19 Pandemic. Sex Transm Dis. 2022; 49:851–854.

14. Dinenno EA, Delaney KP, Pitasi MA, MacGowan R, Miles G, Dailey A, et al. HIV Testing Before and During the COVID-19 Pandemic — United States, 2019–2020. MMWR Morb Mortal Wkly Rep 2022: 71:820–824.

15. Patel D, Williams WO, Wright C, Taylor-Aidoo N, Song W, Marandet A, et al. HIV Testing Services Outcomes in CDC-Funded Health Departments During COVID-19. J Acquir Immune Defic Syndr 2022; 91:117–121

16. Pyra M, Schafer T, Rusie L, Houlberg M, Thompson HM, Hazra A. Temporary changes in STI & HIV testing & diagnoses across different phases of the COVID-19 pandemic, Chicago IL. Front Reprod Health 2023; 5:1072700.

17. Menza TW, Zlot AI, Garai J, Humphrey S, Ferrer J. The Impact of the SARS-CoV-2 Pandemic on Human Immunodeficiency Virus and Bacterial Sexually Transmitted Infection Testing and Diagnosis in Oregon. Sex Transm Dis 2021; 48:E59–E63.

18. Hoover KW, Zhu W, Gant ZC, Delaney KP, Wiener J, Carnes N, et al. HIV Services and Outcomes During the COVID-19 Pandemic-United States, 2019-2021. MMWR Morb Mortal Wkly Rep 2022: 71:1505–1510.

19. Kaiser Family Foundation. Health Insurance Coverage of the Total Population. Kaiser Family Foundation. https://www.kff.org/state-health-policy-data/state-indicator/total-population/?currentTimeframe=0&selectedDistributions=medicaid&sortModel=%7B%22colId%22:%22Location%22,%22sort%22:%22asc%22%7D. [Accessed 14 November 2025]

20. Fultz SL, Skanderson M, Mole LA, Gandhi N, Bryant K, Crystal S, et al. Development and Verification of a ‘Virtual’ Cohort Using the National VA Health Information System. Med Care 2006; 44:S25–30.

21. Chronic Conditions Data Warehouse. Other Chronic Health, Mental Health, and Potentially Disabling Conditions. https://www.tandfonline.com/doi/full/10.1080/09540121.2024.2383901#d1e264. [Accessed 20 August 2025]

22. Pew Research Center. Sharp Divisions on Vote Counts, as Biden Gets High Marks for His Post-Election Conduct. 2020. https://www.pewresearch.org/politics/wp-content/uploads/sites/4/2020/11/PP_2020.11.20_Post-Election-Views_FINAL.pdf [Accessed 10 February 2025].

23. Naing NN. Easy Way to Learn Standardization: Direct and Indirect Methods. Malays J Med Sci 2000; 7:10–5.

24. Brodersen KH, Gallusser F, Koehler J, Remy N, Scott SL. Inferring Causal Impact using Bayesian Structural Time-series Models. Annals of Applied Statistics 2015; 9:247–274.

25. Li X, Li Y, Xu S, Wang P, Hu M, Li H, et al. Evaluation of the impact of COVID-19 on hepatitis B in Henan Province and its epidemic trend based on Bayesian structured time series model. BMC Public Health 2025; 25:1312.

26. Feroze N, Abbas K, Noor F, Ali A. Analysis and forecasts for trends of COVID-19 in Pakistan using Bayesian models. PeerJ. 2021; 7:e11537.

27. Scott SL. BSTS: Bayesian Structural Time Series. https://cran.r-project.org/web/packages/bsts/index.html [Accessed 3 March 2025].

28. Wickham H. Ggplot2: Elegant Graphics for Data Analysis. New York: Springer-Verlag; 2016.

29. Henny KD, Zhu W, Huang YA, Townes A, Delaney KP, Hoover KW. HIV Testing Trends Among Persons with Commercial Insurance or Medicaid-United States, 2014-2019. *MMWR Morb Mortal Wkly Rep* 2021; 70:905-909.

30. Centers for Disease Control and Prevention. Screening and Testing for HIV, Viral Hepatitis, STD & Tuberculosis in Pregnancy, 2024. https://www.cdc.gov/pregnancy-hiv-std-tb-hepatitis/php/screening/index.html [Accessed 24 November 2025].

31. Centers for Disease Control and Prevention. Estimated HIV Incidence and Prevalence in the United States, 2018–2022. https://www.cdc.gov/hiv-data/nhss/estimated-hiv-incidence-and-prevalence.html [Accessed 23 November 2025]

32. Gai Y, Marthinsen, J. Medicaid expansion, HIV testing, and HIV-related risk behaviors in the United States, 2010-2017. Am J Public Health 2019; 109:1404–1412.

33. Song S, Kucik JE. Trends in the Impact of Medicaid Expansion on the Use of Clinical Preventive Services. Am J Prev Med 2022; 62:752–762.

34. Kaiser Family Foundation. Status of State Medicaid Expansion Decisions, 2025 https://www.kff.org/medicaid/status-of-state-medicaid-expansion-decisions/ [Accessed 3 January 2026].

35. Mude W, Mwengyango H, Preston R, O’Mullan C, Vaughan G, Jones G. HIV Testing Disruptions and Service Adaptations During the COVID-19 Pandemic: A Systematic Literature Review. AIDS Behav 2024; 28:186–200.

36. Lyu W, Wehby GL. Effects of the Families First Coronavirus Response Act on Coverage Continuity and Access for Medicaid Beneficiaries. Inquiry 2024; 61: 469580241282052.

37. Sugar S, Peters C, De Lew N, Sommers BD. Medicaid Churning and Continuity of Care: Evidence and Policy Considerations Before and After the COVID-19 Pandemic, 2021. Medicaid Churning and Continuity of Care: Evidence and Policy Considerations Before and After the COVID-19 Pandemic [Accessed 5 January 2026]

