## Supplemental Tables and Figures for "Characterizing the impact of the COVID-19 pandemic on HIV testing among Medicaid beneficiaries"

**Supplementary Table 1**. List of codes used in the study

| Variable | ICD-9 code[a] | ICD-10 Code[a] | CPT/HCPCS |
|---|---|---|---|
| **HIV** | 042, 079.53, 795.71, V08 | B20, B97.35, R75, Z21 | |
| **HIV testing** | | Z11.4 | 86311, 86312, 86314, 86689, 86701, 86702, 86703, 87389, 87390, 87391, 87534, 87535, 87536, 87537, 87538, 87539, G0432, G0433, G0435, G0475, S3645 |

[a] The ICD-9 codes for HIV diagnosis were used purely for excluding beneficiaries with HIV diagnosis at baseline while the ICD-10 codes were used for exclusion if the diagnosis is on or before baseline and determination of incidence of infection if the diagnosis date is after baseline.

**Supplementary Table 2**. Distribution of the follow-up time by baseline year

| Baseline Year | Median [Interquartile range] |
|---|---|
| Overall | 1.50 [0.57, 3.16] |
| 2016 | 3.08 [1.08, 5.50] |
| 2017 | 1.66 [0.58, 4.16] |
| 2018 | 1.58 [0.58, 3.42] |
| 2019 | 2.16 [0.58, 2.58] |
| 2020 | 1.50 [1.25, 1.58] |
| 2021 | 0.50 [0.33, 0.75] |

**Supplementary Table 3**. Comparison of observed and predicted values of HIV testing, HIV diagnosis, and test positivity during the pandemic period

| | Relative effect in % (95% CI) | Posterior tail-area probability p |
|---|---|---|
| **HIV Testing** | | |
| Overall | -8.1 (-12.5, -3.4) | 0.005 |
| Female | | |
| 18 - <35 | -5.1 (-9.2, -0.3) | 0.021 |
| 35 - <50 | -8.3 (-13.3, -2.3) | 0.012 |
| 50 - <65 | -11.4 (-21.5, -0.3) | 0.026 |
| Male | | |
| 18 - <35 | -18.1 (-22.3, -13.8) | <0.001 |
| 35 - <50 | -14.6 (-20.6, -7.3) | 0.004 |
| 50 - <65 | -11.1 (-18.9, -4.5) | 0.006 |
| Black, non-Hispanic | | |
| 18 - <35 | -5.3 (-9.3, -0.3) | 0.022 |
| 35 - <50 | -8.5 (-15.1, -0.5) | 0.022 |
| 50 - <65 | -12.2 (-23.8, -0.5) | 0.027 |
| White, non-Hispanic | | |
| 18 - <35 | -7.3 (-12.4, -0.1) | 0.025 |
| 35 - <50 | -12.2 (-18.3, -3.6) | 0.011 |
| 50 - <65 | -10.4 (-19.6, -2.1) | 0.015 |
| COVID-19 restriction | | |
| Least | -4.8 (-12.1, 3.9) | 0.074 |
| Less | -8.0 (-12.1, -4.0) | 0.004 |
| Most | -9.3 (-15.4, 1.4) | 0.032 |
| **HIV infection** | | |
| Overall | 2.2 (-5.1, 9.4) | 0.201 |
| Female | | |
| 18 - <35 | 12.6 (-11.5, 76.3) | 0.189 |
| 35 - <49 | 13.8 (-11.7, 70.0) | 0.159 |
| 50 - <65 | -10.1 (-37.3, 20.7) | 0.189 |
| Male | | |
| 18 - <35 | 2.9 (-4.5,10.5) | 0.154 |
| 35 - <49 | -0.7 (-16.2,12.0) | 0.483 |
| 50 - <65 | 2.6 (-6.6,14.8) | 0.258 |
| Black, non-Hispanic | | |
| 18 - <35 | 4.9 (-2.8,16.3) | 0.090 |
| 35 - <49 | 2.4 (-15.2,16.6) | 0.306 |
| 50 - <65 | 2.7 (-10.9,16.9) | 0.288 |
| White, non-Hispanic | | |
| 18 - <35 | 8.5 (-9.9, 46.0) | 0.206 |
| 35 - <49 | 1.3 (-19.2, 30.8) | 0.499 |
| 50 - <65 | 3.4 (-10.0,27.4) | 0.361 |
| COVID-19 restriction | | |
| Least | 1.0 (-15.1,23.1) | 0.498 |
| Less | 5.1 (-3.5,14.2) | 0.080 |
| Most | -1.1 (-18.5,17.0) | 0.380 |
| **Test positivity** | | |
| Overall | 6.2 (-14.5, 18.8) | 0.121 |
| Female | 0.3 (-16.8, 21.4) | 0.492 |
| Male | 19.7 (-95.5, 38.2) | 0.193 |
| Black, non-Hispanic | 4.0 (-41.3, 26.2) | 0.212 |
| White, non-Hispanic | 9.0 (-19.5, 31.9) | 0.151 |
| COVID-19 restriction | | |
| Least | -7.8 (-54.8, 22.8) | 0.421 |
| Less | 12.2 (-27.7, 31.2) | 0.107 |
| Most | 10.4 (-13.2, 30.7) | 0.468 |

Note: CI – Confidence interval

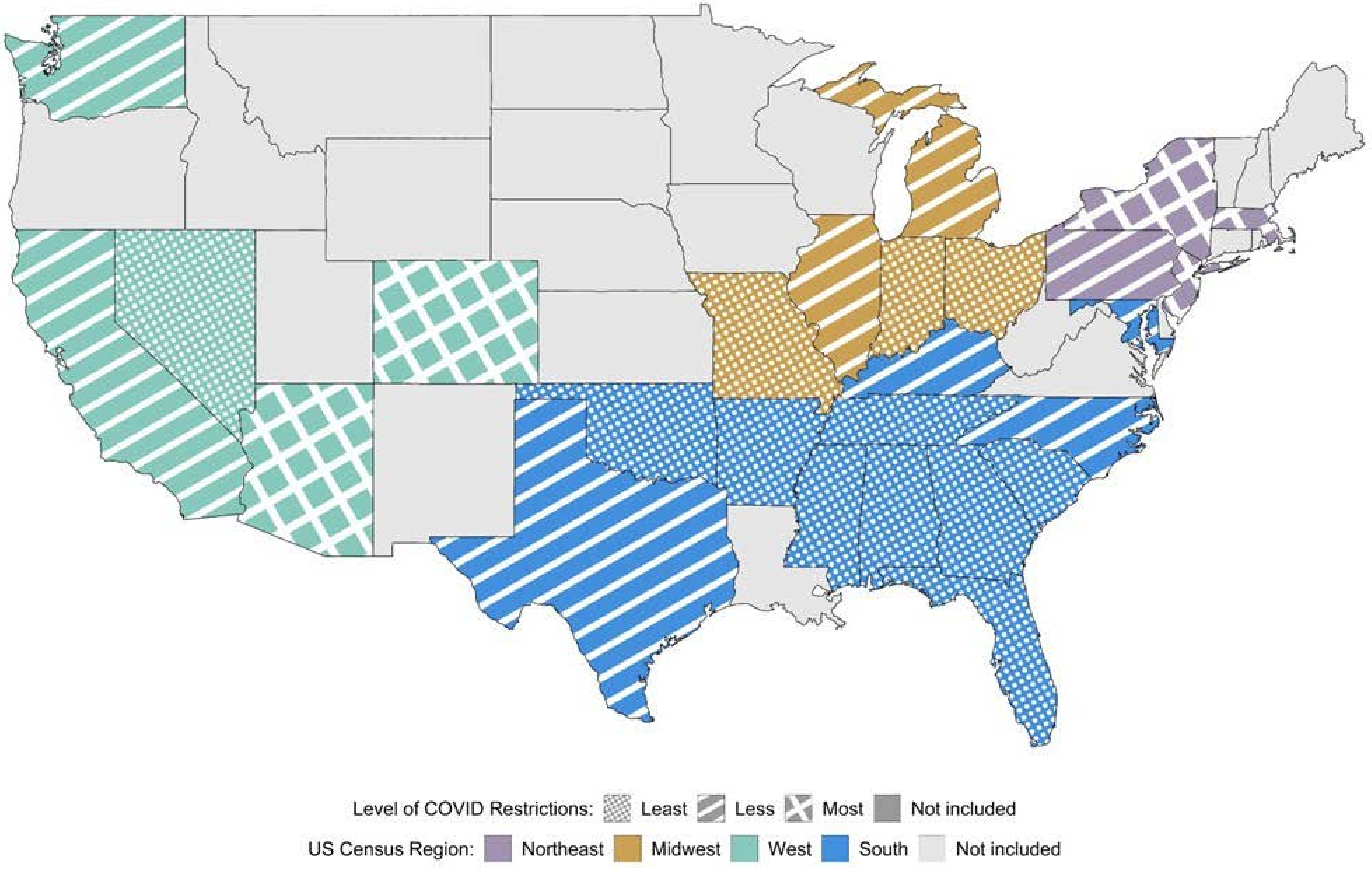


***Supplementary Figure 1***. US states included in the study by level of COVID-19 restrictions and census region

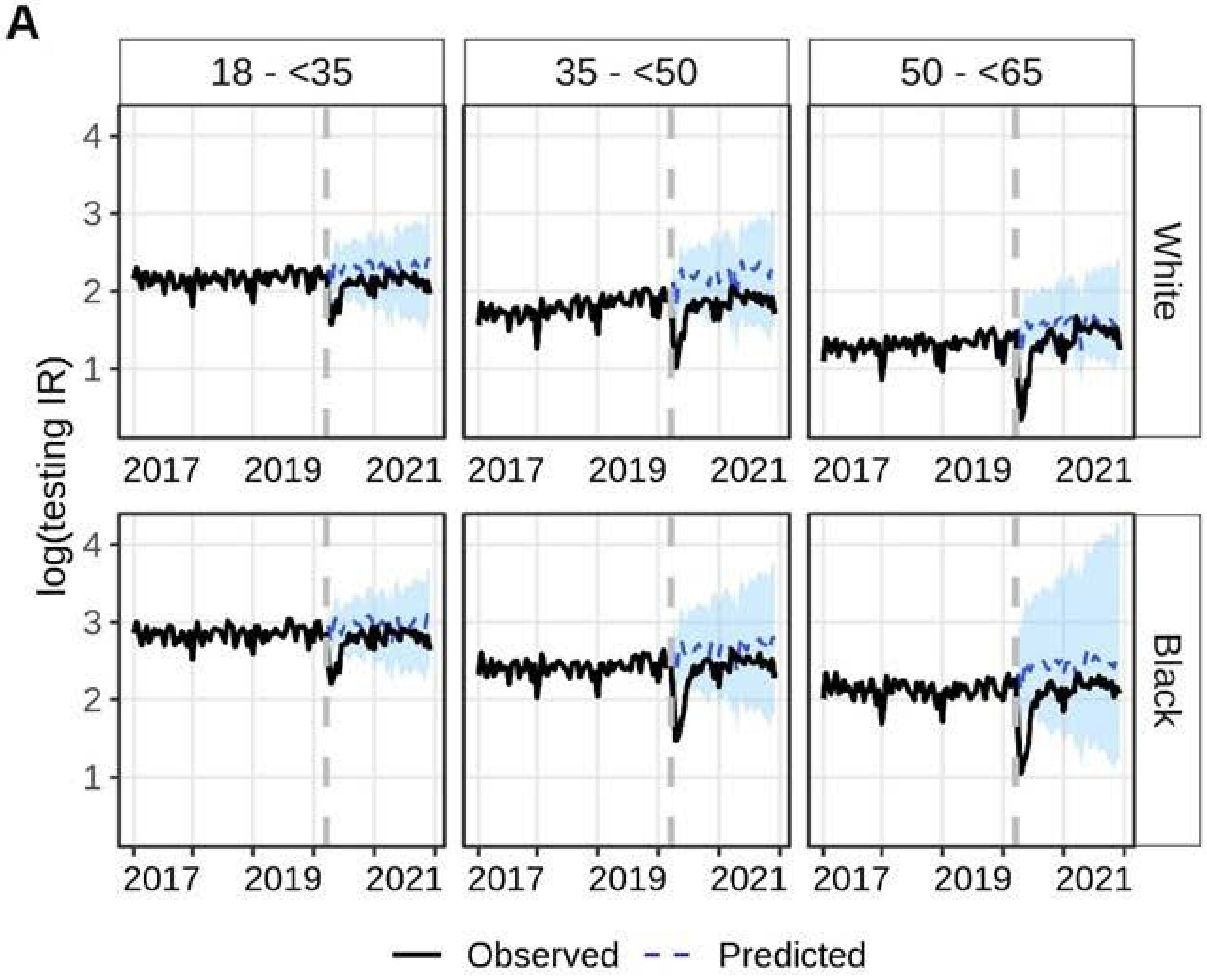


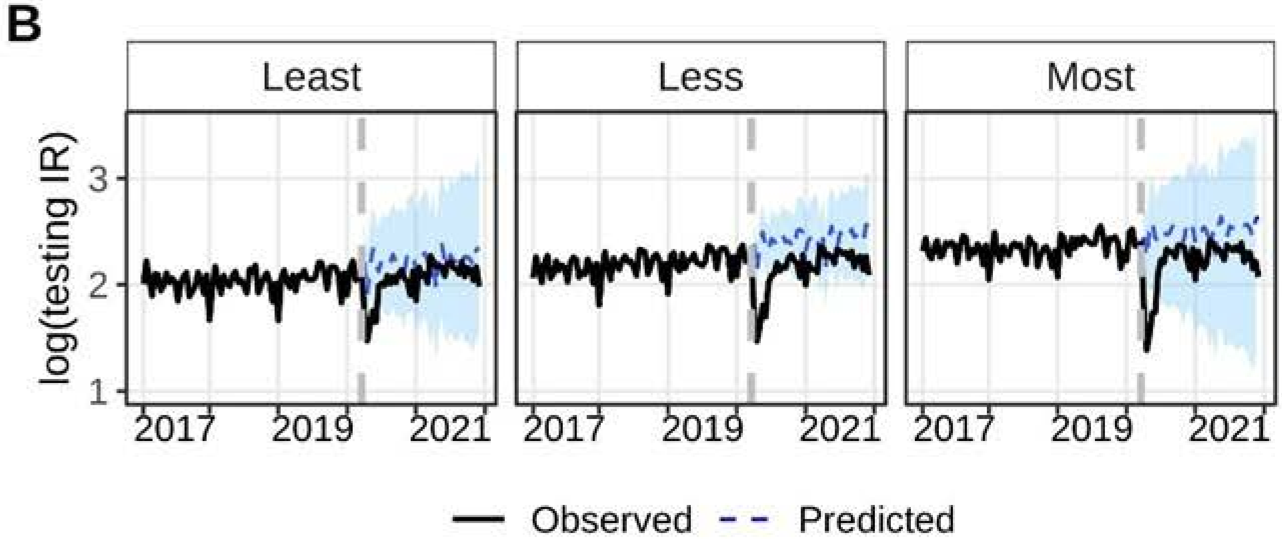


***Supplementary Figure 2***. Trend of log(standardized HIV testing incidence rate) from January 2017 to December 2021 stratified by A) age and race, and B) states' COVID-19 restriction policies. The vertical gray dashed line separated the pre-pandemic and pandemic periods. The solid lines represent the observed values while the blue dashed lines with blue 95% credible intervals during the pandemic period represent the predicted values had the pandemic not happened.

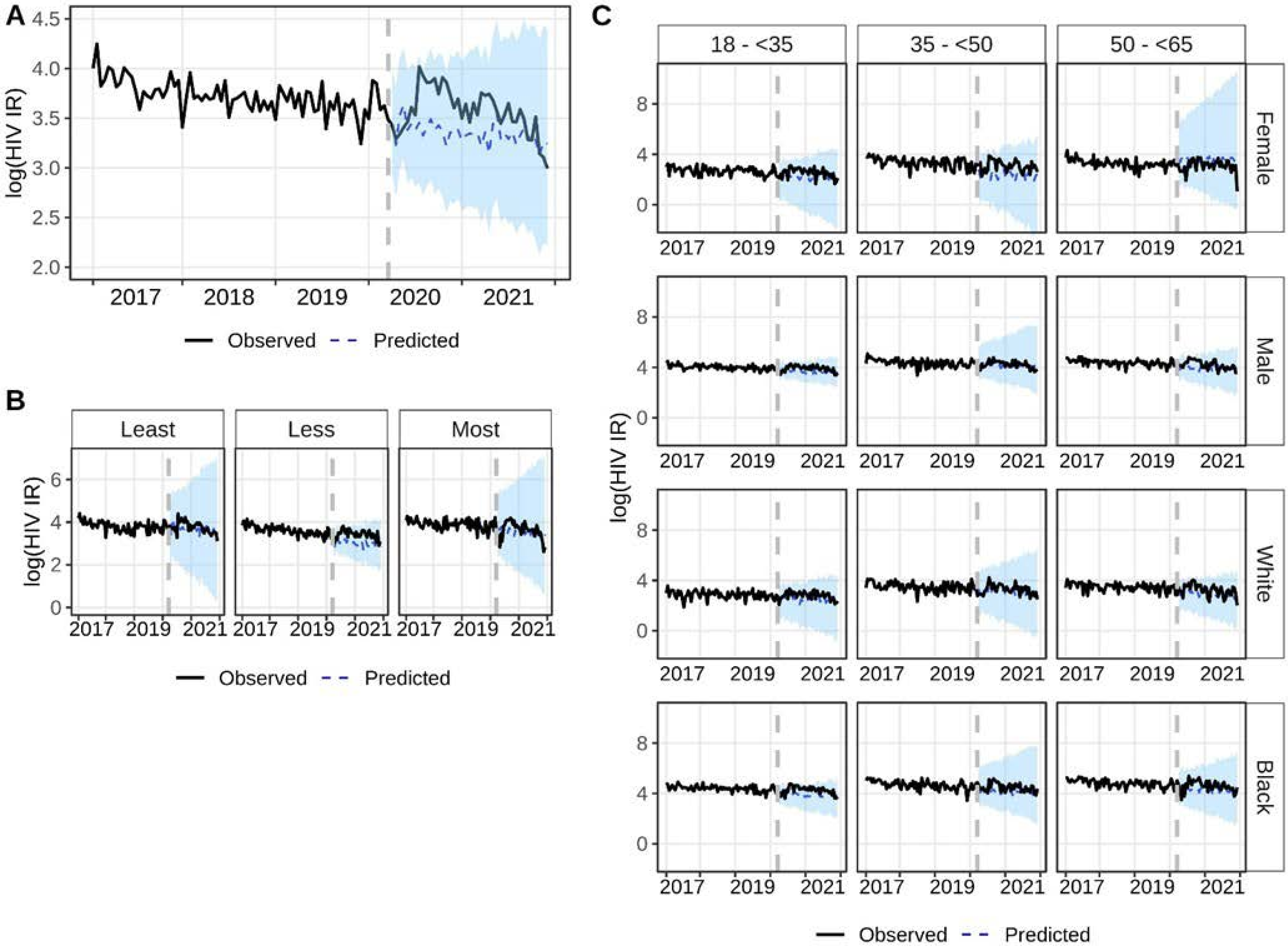


***Supplementary Figure 3***. Trend of log(standardized HIV incidence rate) from January 2017 to December 2021 A) overall, B) by states' COVID-19 restriction policies, and C) by age, sex, and race/ethnicity. The vertical gray dashed line separated the pre-pandemic and pandemic periods. The solid lines represent the observed values while the blue dashed lines with blue 95% credible intervals during the pandemic period represent the predicted values had the pandemic not happened.

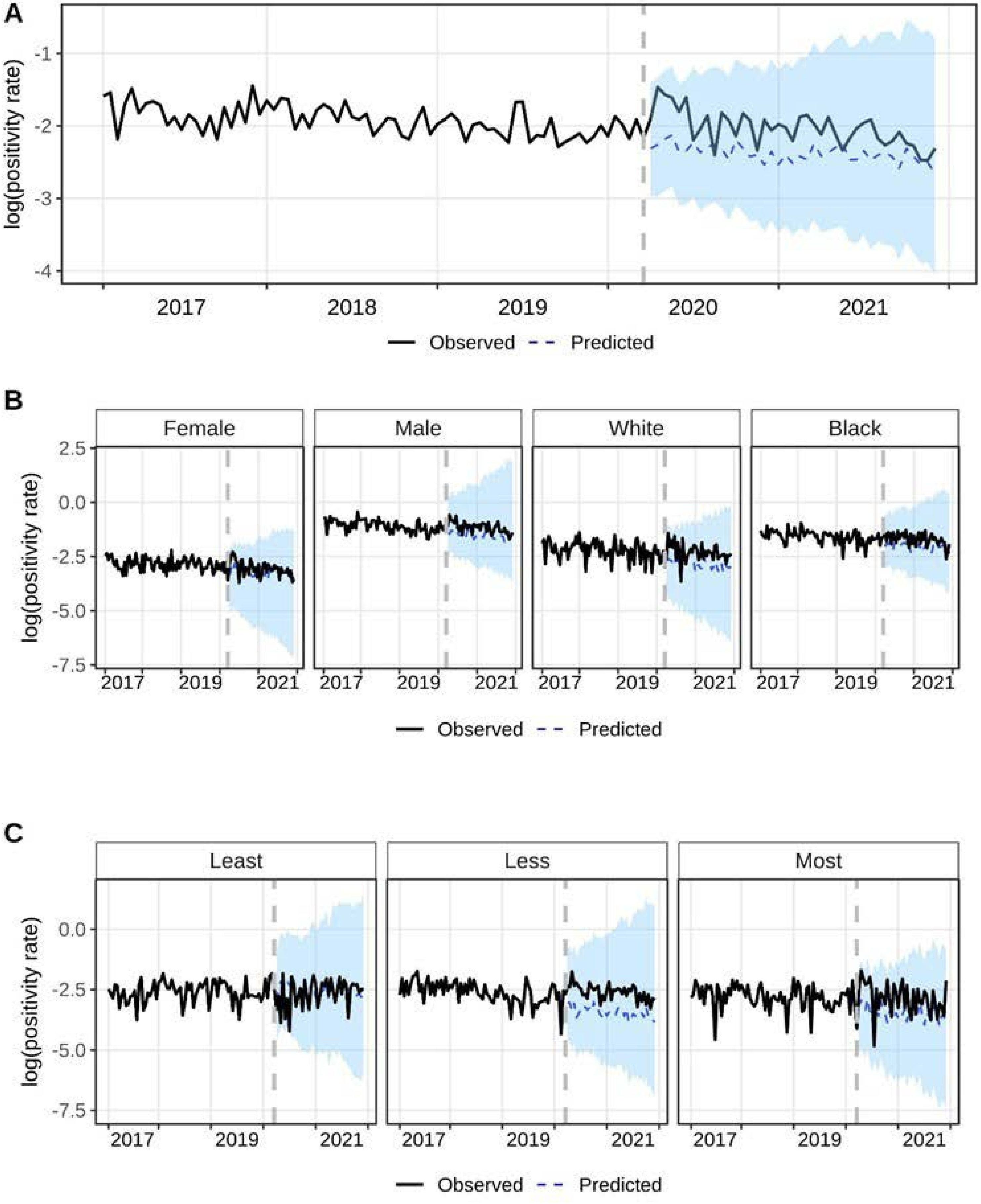


***Supplementary Figure 4***. Trend of log(standardized test positivity rate) from January 2017 to December 2021 A) overall, B) by sex and by race, and C) by states' COVID-19 restriction policies. The vertical gray dashed line separated the pre-pandemic and pandemic periods. The solid lines represent the observed values while the blue dashed lines with blue 95% credible intervals during the pandemic period represent the predicted values had the pandemic not happened.
